# Optimized Machine Learning Algorithms for the Classification and Diagnosis of Sleep Disorders

**DOI:** 10.1101/2025.10.07.25337263

**Authors:** Timothy Oluwapelumi Adeyemi

## Abstract

Sleep disorders, including insomnia and obstructive sleep apnea, affect millions of individuals worldwide but are frequently undetected due to the high cost, limited availability, and complexity of conventional diagnostic tools such as polysomnography. This study presents an interpretable machine learning framework for multi-class sleep disorder classification that utilizes routine clinical and lifestyle data, offering an accessible, data-driven approach to screening. The dataset was preprocessed and balanced using SMOTE-ENN, and three ensemble models, Random Forest, XGBoost, and LightGBM, were systematically trained and optimized. Among these, LightGBM demonstrated superior performance, achieving a test accuracy, precision, and recall of 0.9835 above with a minimal train–test accuracy gap of 0.0105, indicating strong generalization and limited overfitting. Feature importance analysis identified systolic blood pressure and BMI category as the most influential predictors, consistent with established clinical knowledge. To enable practical application, the optimized model was deployed through a lightweight Flask-based web application, providing real-time, point-of-care predictions without requiring specialized equipment. The findings demonstrate that well-tuned tree-based ensemble models can deliver accurate, interpretable, and clinically actionable tools for the early detection of sleep disorders, supporting timely intervention and expanding access to sleep health management across diverse healthcare settings.

## 1 INTRODUCTION

Sleep is not merely a state of rest, but an active and restorative process essential for cognitive renewal, emotional regulation, and physical health (Nayana, 2025). Disorders such as insomnia and sleep apnea remain among the most critical yet frequently underdiagnosed conditions, with profound implications that include impaired daily performance, increased cardiovascular and metabolic risk, and decreased quality of life (Niu et al., 2025). Recent surveys indicate that more than 40% of adults report dissatisfaction with their sleep, with stress, occupational demands, and lifestyle habits emerging as dominant contributing factors (Triches et al., 2025).

Historically, clinical diagnosis depended on polysomnography (PSG), a laboratory-based examination that combines electroencephalographic, respiratory, cardiac, and movement signals (Kathrine et al., 2025). Despite its diagnostic accuracy, PSG is expensive, time-consuming, and unsuitable for population-level screening. Furthermore, its interpretation is subject to inter-rater variability, which limits repeatability. The growing availability of multimodal health data, including wearable sensors and electronic health records, underscores the need for objective, scalable, and automated diagnostic frameworks that utilize accessible biomarkers such as heart rate variability, blood pressure, physical activity, perceived stress, body composition, and demographic covariates (Elfouly and Alouani, 2025).

Machine learning (ML) offers a transformative opportunity in this field. By identifying nonlinear interactions and hidden patterns in various data streams, ML models can classify individuals into clinically relevant subgroups, allowing precision therapies (Tiwari et al., 2025). However, much of the current literature remains limited: many studies focus narrowly on single-modality signals (e.g., EEG or actigraphy) (Gramkow et al., 2025; Krauss et al., 2025; Bjarke Mikkelsen et al., 2025), overlooking the synergistic effects of psychosocial, physiological, and behavioral factors, such as how chronic stress influences the relationship between BMI and sleep problems. In addition, there is often little comparative benchmarking among algorithms, and limited efforts are made to turn successful models into practical clinical tools.

To close these gaps, this study proposes a robust end-to-end machine learning system to classify three states of sleep: no disorder, insomnia, and sleep apnea. Physiological, demographic, lifestyle, and mental health indicators are integrated through systematic preprocessing, feature engineering, and model optimization. Three advanced ensemble methods, Random Forest, XGBoost, and LightGBM, are extensively tested under similar experimental conditions to ensure a fair comparison. Our approach improves the precision of diagnosis, reduces the reliance on subjective clinical interpretation, and promotes early and accessible intervention.

The key contributions of this work are threefold:

1. A comprehensive review of existing research on machine learning for the classification of sleep disorders, focusing on key challenges and limitations.
2. A comparative evaluation of traditional machine learning models and deep learning architectures for an accurate classification of sleep disorders.
3. A prototype web application that integrates the developed models to offer accessible and automated diagnostic support for clinicians and patients.

The remainder of this paper is structured as follows. Section 2 reviews previous studies; Section 3 describes the data set and methodology; Section 4 presents and discusses the experimental findings; and Section 5 provides concluding insights and recommendations.

## 2 RELATED WORKS

Recent advances in machine learning (ML) and deep learning (DL) have improved the automated classification of sleep disorders, shifting from traditional signal-based detection to multimodal and clinically interpretable frameworks. This section reviews existing approaches, focusing on their methodologies, strengths, and limitations.

Detecting sleep apnea is difficult due to the frequent underdiagnosis of hypopnea events. (Yook et al., 2024) proposed a deep learning pipeline that transforms physiological signals, nasal airflow, SpO_2_, and ECG into Morlet wavelet scalograms, which are then processed using an Xception-based CNN. The model reached 94% accuracy in diagnosing apnea, hypopnea, and regular breathing, with the highest performance obtained by integrating SpO_2_ and airflow data. Event-level predictions were combined with demographic characteristics using an SVM, resulting in 99% precision for OSA screening and 93% for severity staging. Despite robust validation, performance was worse in patients with hypopnea-dominant conditions, and limitations included single-center data and the exclusion of central apnea.

In addition to direct disorder classification, (Li et al., 2024) investigates the prediction of psychiatric comorbidities within the OSAHS population using the NHANES database. Their comparative analysis of logistic regression, LASSO, and random forest models identifies key predictors of depression, including female sex, obesity, poor self-rated health, and severe OSAHS. Logistic regression demonstrates the most significant clinical utility due to its interpretability and balanced performance (AUC = 0.746). The development of a clinician-friendly nomogram, combined with the use of decision curve analysis, further supports its application for point-of-care risk stratification. However, the study’s reliance on self-reported snoring rather than PSG-confirmed AHI and its cross-sectional design limit causal inference.

(Rahman et al., 2025) establishes a methodologically rigorous benchmark for classifying common sleep disorders (none, insomnia, sleep apnea) using the publicly available Sleep Health and Lifestyle Dataset. Their pipeline incorporates SMOTEENN for class balancing, ANOVA for feature significance, and stacking-based feature engineering. Ensemble models, such as Gradient Boosting, CatBoost, and Stacking, achieve a precision of 97.33%, surpassing previous work (e.g., Alshammari’s accuracy of 92.92% with ANN). A lightweight decision tree that uses only two engineered features achieves 96% accuracy with 149-fold faster inference, demonstrating the potential for efficient, high-performance models in practical settings. However, generalizability is limited by the modest sample size of the dataset (n = 374).

Extending the research, (Nyholm et al., 2024) explores sleep disturbances as early signs of dementia in a large longitudinal cohort (SNAC-B, n = 4,175). Using 16 features related to those in a gradient-boosting model, they achieve 92.9% accuracy (AUC = 0.97) in predicting dementia. Key predictors include daytime sleep over two hours and snoring. Although the findings are statistically significant, the small effect size (Pseudo R^2^ = 0.0565) indicates that sleep is only one factor among many that influence the risk of dementia. Differences in the importance of features between models underscore the need for larger and more diverse datasets to improve the reliability of the model. In a related approach, (Kim et al., 2024) bypasses biosignals and directly diagnoses the severity of OSA from routine craniofacial CT scans using a multimodal 3D CNN (AirwayNet-MM-H). By combining structural imaging, clinical metadata, and a preprocessing step that highlights airways, this model achieves 87.6% accuracy in predicting four severity classes of OSA, with an AUC of 0.910 for moderate to severe OSA. Validation was carried out in three cohorts. This screening method uses existing scans (such as those for sinusitis) to identify undiagnosed OSA. However, reliance on standardized CT protocols and validation in only Asian populations raises questions about the broad applicability of the results.

(de Araújo Costa et al., 2024) address post-diagnostic decision support by proposing CROWM. This novel framework integrates Principal Component Analysis (PCA) with Multicriteria Decision Analysis (MCDA) to objectively rank CPAP devices based on practical criteria, including cost, noise, and weight. By deriving criterion weights from PCA communalities and the MEREC method, CROWM minimizes subjective bias and identifies the AirMini AutoSet as the best-performing device. Robustness is confirmed through sensitivity analysis, yielding high rank consistency across alternative methods (Spearman’s *ρ >* 0.817). Although the exclusion of patient-reported outcomes limits clinical granularity, the approach offers a transparent, data-driven foundation for personalized therapy selection.

Alternative signal modalities are increasingly being explored. (Padovano et al., 2025) applied convolutional neural networks (CNNs) to nonthresholded recurrence plots derived from time series of heart rate variability (HRV). This method captures complex nonlinear patterns that traditional recurrence quantification analysis might miss. Their AlexNet-based model achieved 74.7% accuracy in obstructive sleep apnea (OSA) detection during rigorous external validation, surpassing results from thresholded recurrence plots and standard machine learning techniques. These findings suggest that deep learning can leverage the inherent geometric features of physiological dynamics for diagnostic purposes.

Meanwhile, (Dang et al., 2024) proposes a non-invasive acoustic screening method. It employs a 1D CNN to classify snore sounds from nocturnal audio, achieving 98.65% precision and 95.1% recall. The system is optimized for mobile deployment through TensorFlow Lite and facilitates real-time home monitoring. However, its clinical validity remains unproven, as it assumes that snore acoustics alone can distinguish between benign snoring and pathological apnea, a critical gap that requires PSG correlation.

(Alshammari, 2024) conduct a systematic comparison of five machine learning models in the Sleep Health and Lifestyle Dataset, incorporating hyperparameter tuning based on generic and feature selection. The GA-optimized artificial neural network (ANN) achieves 92.92% accuracy, indicating that deep learning models significantly benefit from metaheuristic optimization in this context. Despite technical rigor, the study is limited by a lack of clinical interpretability and the non-clinical nature of the dataset.

Population-level insights emerge from a study by (Al-Mamun et al., 2024) of 1,496 Bangladeshi university entrance exam candidates, revealing alarmingly high prevalence rates of insomnia (25.5%) and abnormal sleep duration (62.9%), both of which are associated with academic stress. Although the CatBoost model achieves 73.46% accuracy, its limited discriminative capacity (AUC *leq*0.62) undermines diagnostic reliability. GIS-based spatial analysis reveals significant regional disparities, indicating that socio-environmental factors play a crucial role in sleep health, with important implications for targeted public health interventions.

(Varghese et al., 2025) presents ApneaFormer, a transformer-based model for OSA detection from raw ECG, validated on both healthy (PhysioNet) and complex clinical (OSASUD) cohorts. By integrating dynamical system features (e.g., Lyapunov exponent) and SHAP explainability, they ground predictions in physiological mechanisms. Their successful transfer learning and patient-specific personalization strategies represent a significant step toward the deployment of robust models in heterogeneous clinical settings.

Finally, returning to the fundamental biosignal in sleep medicine, (Yousefi and Rahimi, 2025) focuses on electroencephalography (EEG), the gold standard for sleep staging, which differentiates between healthy and disordered sleep using long short-term memory (LSTM) networks. Using the Sleep-EDF database of 197 full-night recordings, their 12-layer LSTM process preprocessed EEG features extracted from 10-second windows, achieving 93.3% accuracy in binary classification. Through a fusion strategy based on majority vote in multiple model runs, they further improve performance to 95%. The study presents a transparent and reproducible pipeline, covering notch and Butterworth filtering, feature engineering, and hyperparameter tuning (125 epochs, 200 hidden units, learning rate = 0.01), which demonstrates the ability of LSTM’s to capture the temporal dynamics present in EEG for disorder detection. While the binary task simplifies clinical complexity compared to multi-class staging, the authors pragmatically acknowledge the model’s computational demands and suggest meta-heuristic optimization as a way to enhance clinical feasibility.

Collectively, these studies demonstrate a trend toward combining innovation and clinical pragmatism via multimodal data, interpretability, efficient models, and innovative sources. Limited datasets, demographic biases, and the gap between accuracy and clinical actionability are also ongoing issues. This study builds upon previous attempts by focusing on algorithmic optimization to enhance both performance and real-world deployability in sleep diagnostics.

## 3 METHODOLOGY

This section outlines the end-to-end machine learning process used to classify sleep disorders. The framework incorporates data preprocessing, feature engineering, class balance management, model selection, rigorous performance evaluation, and statistical validation. To ensure reproducibility, promote computational efficiency, and uphold methodological rigor, all experiments were implemented in Python with robust scientific libraries, including `NumPy`, `pandas`, `scikit-learn`, `XGBoost`, `LightGBM`, `SciPy`, and `statsmodels`. Figure 1 presents an overview of the whole procedure.

**Figure 1.**
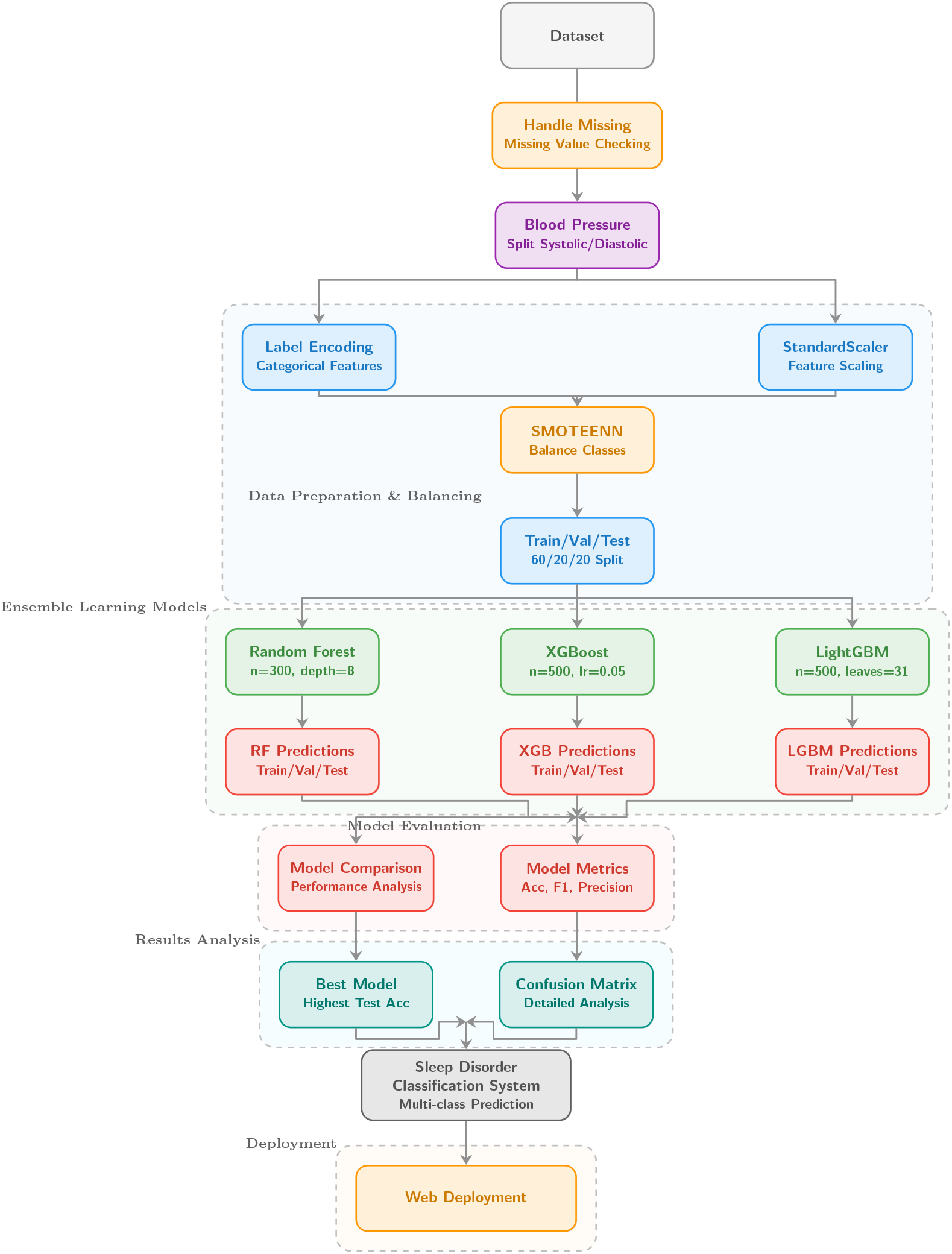
End-to-end pipeline for sleep disorder classification and diagnosis.

### 3.1 Dataset Description

The study uses the publicly accessible Sleep Health and Lifestyle dataset from Kaggle (Tharmalingam, 2024), which includes 374 anonymized records across 13 categories. The dataset consists of both categorical and numerical variables and covers individuals with a wide range of demographic and lifestyle characteristics. The target variable, Sleep Disorder, is a multiclass label that includes ‘None’, ‘Insomnia’, and ‘Sleep Apnea’.

The Person ID variable, which just serves as a unique identifier, was removed from the modeling process because it has no predictive value. Table 2 provides a complete summary of the dataset. Panel A shows categorical distributions, while Panel B offers descriptive statistics (mean, standard deviation, and range) for numerical features.

**Table 1.** Key Studies on Machine Learning for Sleep Disorder Diagnosis.

| Author | Approach Used | Main Results | Why It Matters |
| --- | --- | --- | --- |
| Yook et al. (2024) | Combined Xception CNN on multimodal sleep signals (SpO<sub>2</sub>, ECG) with SVM for severity staging. | Correctly identified 94% of events and 99% of sleep apnea cases, reducing errors in mild or moderate cases. | Blending machine learning with medical guidelines enables highly accurate and practical diagnosis. |
| Rahman et al. (2025) | Applied ensemble models (Gradient Boosting, CatBoost, Stacking) with SMOTE-ENN resampling and advanced feature engineering. | Achieved 97.3% accuracy. A simpler version achieved 96% accuracy while being 149 times faster. | Highlights the balance between high accuracy and efficiency, making it more suitable for real-world use. |
| Yousefi and Rahimi (2025) | Deployed 12-layer LSTM architecture on preprocessed EEG signals with majority-vote fusion. | Improved accuracy from 93.3% to 95% in separating healthy sleep from disordered sleep. | Demonstrates the power of time-based analysis for detecting sleep problems in clinical settings. |
| Varghese et al. (2025) | Introduced Transformer-based ApneaFormer on raw ECG with dynamical features, SHAP explainability, and transfer learning. | Reached strong performance (F1 = 0.914) across different datasets and adapted well to individual patients. | Combines accuracy, adaptability, and interpretability, key for clinical acceptance. |
| Alshammari (2024) | Optimized Artificial Neural Network (ANN), SVM, and Random Forest using a Genetic Algorithm (GA). | Found that the optimized neural network achieved 92.9% accuracy, the highest among the tested models. | Shows that fine-tuning models with optimization techniques can improve performance in diagnosing sleep disorders. |
| Kim et al. (2024) | Developed multimodal 3D CNN (AirwayNet-MM-H) combining craniofacial CT scans with clinical metadata. | Achieved 87.6% accuracy in distinguishing sleep apnea severity, validated across groups. | Provides a way to detect sleep apnea without expensive sleep studies, using routine medical scans. |
| Padovano et al. (2025) | Applied AlexNet CNN to unthresholded HRV recurrence plots derived from heart dynamics. | Achieved 74.7% accuracy under strict external tests, outperforming traditional methods. | Shows potential of end-to-end learning from heart signals for detecting sleep apnea. |
| Li et al. (2024) | Utilized Logistic Regression with LASSO and Random Forest for feature selection on NHANES survey data. | Achieved an AUC of 0.746 and identified key risk factors like depression in sleep apnea patients. | Offers a simpler, interpretable approach to identify risks and support other diagnostic tools. |

**Table 2.**
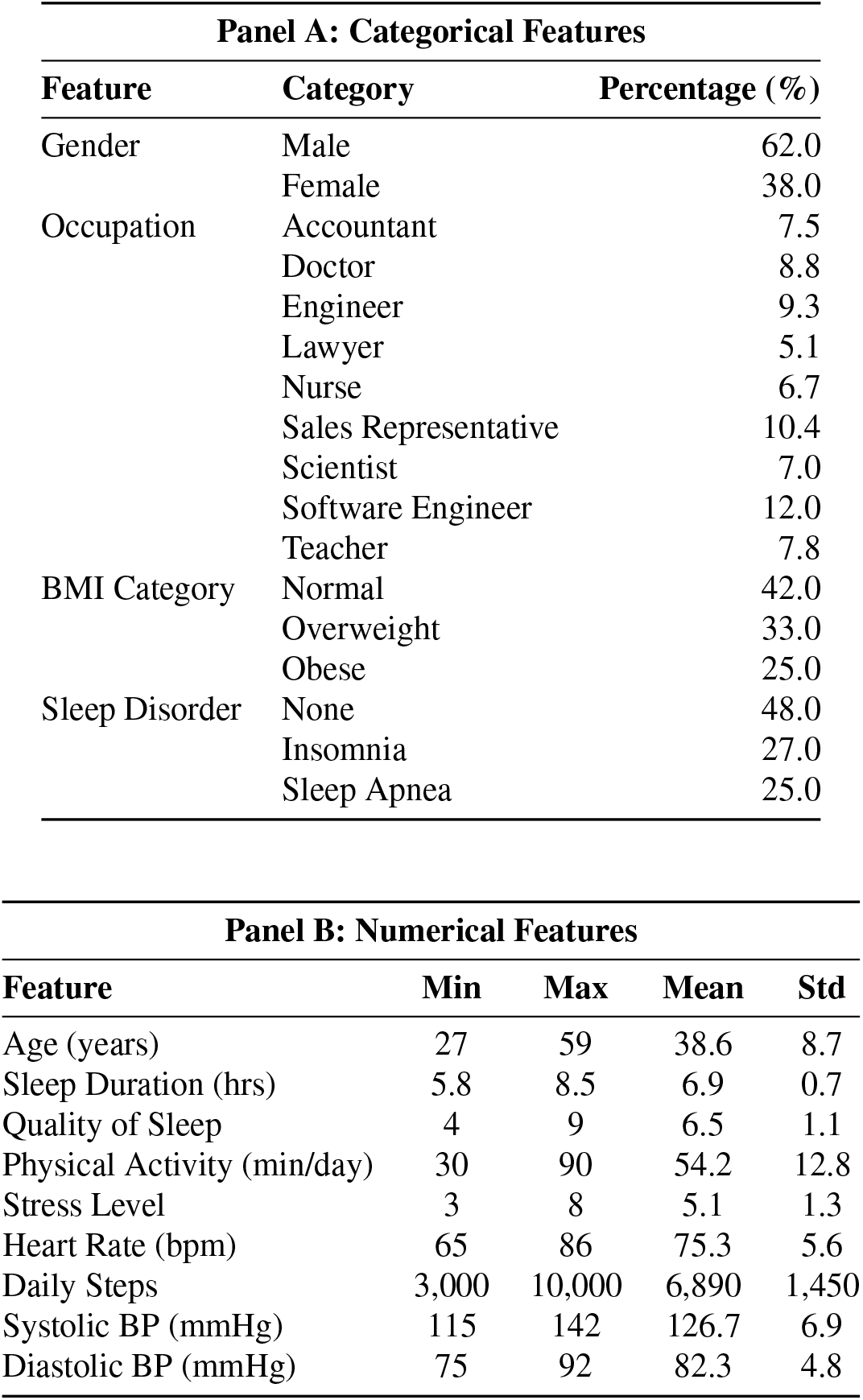
Summary of dataset features: categorical distributions (Panel A) and numerical statistics (Panel B).

| Panel A: Categorical Features |  |  |
| --- | --- | --- |
| Feature | Category | Percentage (%) |
| Gender | Male | 62.0 |
|  | Female | 38.0 |
| Occupation | Accountant | 7.5 |
|  | Doctor | 8.8 |
|  | Engineer | 9.3 |
|  | Lawyer | 5.1 |
|  | Nurse | 6.7 |
|  | Sales Representative | 10.4 |
|  | Scientist | 7.0 |
|  | Software Engineer | 12.0 |
|  | Teacher | 7.8 |
| BMI Category | Normal | 42.0 |
|  | Overweight | 33.0 |
|  | Obese | 25.0 |
| Sleep Disorder | None | 48.0 |
|  | Insomnia | 27.0 |
|  | Sleep Apnea | 25.0 |

| Panel B: Numerical Features |  |  |  |  |
| --- | --- | --- | --- | --- |
| Feature | Min | Max | Mean | Std |
| Age (years) | 27 | 59 | 38.6 | 8.7 |
| Sleep Duration (hrs) | 5.8 | 8.5 | 6.9 | 0.7 |
| Quality of Sleep | 4 | 9 | 6.5 | 1.1 |
| Physical Activity (min/day) | 30 | 90 | 54.2 | 12.8 |
| Stress Level | 3 | 8 | 5.1 | 1.3 |
| Heart Rate (bpm) | 65 | 86 | 75.3 | 5.6 |
| Daily Steps | 3,000 | 10,000 | 6,890 | 1,450 |
| Systolic BP (mmHg) | 115 | 142 | 126.7 | 6.9 |
| Diastolic BP (mmHg) | 75 | 92 | 82.3 | 4.8 |

### 3.2 Data Preprocessing

Initial data validation confirmed the absence of missing values and duplicate records, thereby maintaining dataset integrity. The initial `Blood Pressure` field, saved as a string (e.g., “120/80”), was parsed into two numerical features: `SystolicBP` and `DiastolicBP`. All categorical predictors were transformed into numerical representations using label encoding. The target variable `Sleep Disorder` was encoded as integers: `Insomnia` → 0, `None` → 1, `Sleep Apnea` → 2.

To reduce scale inequalities across features and guarantee fair influence during model improvement, all input variables were normalized using z-score normalization:

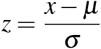

*µ* and *σ* represent the feature-specific mean and standard deviation determined on the training set.

### 3.3 Class Balancing and Data Partitioning

The original dataset exhibited considerable class imbalance (Table 3). To address this, the hybrid SMOTEENN technique was applied, combining the Synthetic Minority Oversampling Technique (SMOTE) with Edited Nearest Neighbors (ENN) to generate synthetic minority samples while removing noisy or borderline instances from the majority class. This approach enhanced class separability and reduced the risk of overfitting.

**Table 3.** Class distribution before and after SMOTE-ENN balancing.

| Class | Before SMOTE-ENN | After SMOTE-ENN |
| --- | --- | --- |
| None | 219 | 168 |
| Insomnia | 77 | 190 |
| Sleep Apnea | 78 | 187 |
| **Total** | **374** | **545** |

Following resampling, the dataset was expanded to *N* = 545 and partitioned into training, validation, and test subsets using stratified random sampling to preserve the proportional representation of each sleep disorder class across all splits. The final allocation followed a 60:20:20 ratio, resulting in 327 training, 109 validation, and 109 test samples. Stratification ensured rigorous evaluation and improved generalizability, particularly for minority classes.

### 3.4 Model Development

The study assessed three ensemble-based classifiers known for their effective performance on structured tabular data: Light Gradient Boosting Machine (LightGBM), a histogram-based boosting algorithm optimized for speed and memory efficiency; Extreme Gradient Boosting (XGBoost), a regularized gradient boosting framework; and Random Forest, an averaging ensemble of decorrelated decision trees. Using cross-validation on the validation set and employing empirical best practices, hyperparameters were optimized through repeated testing. Table 4 displays the final model settings, which were created to balance regularization, computational efficiency, and prediction accuracy.

**Table 4.** Hyperparameter configurations for ensemble classifiers.

| Parameter | RF | XGBoost | LightGBM | Rationale |
| --- | --- | --- | --- | --- |
| Number of estimators | 300 | 500 | 500 | Ensures convergence without excessive cost |
| Max depth | 8 | 6 | 6 | Controls complexity and avoids overfitting |
| Learning rate | – | 0.05 | 0.05 | Enables stable gradient updates |
| Subsample | – | 0.8 | 0.8 | Adds randomness to reduce variance |
| Colsample.bytree | – | 0.8 | 0.8 | Promotes feature diversity |
| Num leaves | – | – | 32 | Limits tree growth in LightGBM |
| Min samples split | 10 | – | – | Avoids splits on very small subsets |
| Min samples leaf | 5 | – | – | Regularizes terminal nodes |
| Min data in leaf | – | – | 20 | Ensures enough samples per leaf |
| L1/L2 regularization ($\alpha/\lambda$) | – | 1/1 | 1/1 | Penalizes complexity |
| Max features | sqrt | – | – | Random feature selection per split |
| Random state | 42 | 42 | 42 | Ensures reproducibility |

### 3.5 Evaluation Metrics

This study evaluates machine learning algorithms for categorizing sleep problems. To address class imbalance, four measures were employed: accuracy, precision, recall, and weighted *F*_1_-score, along with train-test and validation-test gaps to assess generalization and overfitting.

- **Accuracy**: Measures the overall correctness of predictions.

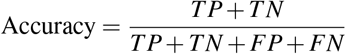
- **Precision**: The proportion of correctly identified positive cases among all predicted positives.

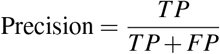
- **Recall (Sensitivity)**: The proportion of actual positive cases correctly identified.

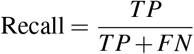
- **Weighted** *F*_1_**-score**: The harmonic mean of precision and recall, averaged across classes using support-based weighting. as:

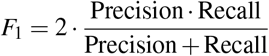

To evaluate generalization and detect potential overfitting, the **train–test accuracy gap** was computed

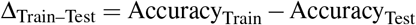

Similarly, the **validation–test accuracy gap** was used to assess stability across unseen data:

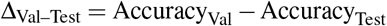

In addition to these numerical measures, confusion matrices and classification reports were generated to capture per-class misclassification patterns. Collectively, these metrics provided a robust and comprehensive evaluation of classification performance and model generalization.

### 3.6 Model Deployment

The best-performing model, selected based on test accuracy, weighted *F*_1_-score, and the smallest generalization gap, was deployed as a lightweight web application utilizing the `Flask` framework. Flask was selected due to its ease of use, modularity, and seamless compatibility with machine learning libraries based on Python.

The trained model was serialized using `joblib` to retain scikit-learn compatibility and facilitate quick loading during inference. To guarantee consistency with training settings, input preprocessing (label encoding, standardization) was duplicated exactly in the deployment environment.

Clinicians or researchers can use a browser to enter patient-specific data (such as demographics, vital signs, and lifestyle measures) into the resultant interface. The backend returns a real-time prediction of the sleep disorder category after preprocessing the inputs and running the model. Adoption in clinical or public health settings is facilitated by this design, which eliminates the requirement for technical competence.

This deployment demonstrates the translational potential of machine learning in sleep medicine by integrating experimental modeling and real-world implementation, facilitating data-driven clinical decision-making, enabling early screening, and reducing diagnostic delays.

## 4 RESULTS AND DISCUSSION

The empirical findings of the experimental study are presented, with an emphasis on the model’s performance, interpretability, and therapeutic applicability. Comparison research was conducted to determine the most effective practical strategy for diagnosing sleep problems, comparing three ensemble classifiers: Random Forest (RF), XGBoost, and LightGBM, based on several assessment criteria. The commentary situates the findings within the larger body of research, balancing their strengths and limitations in terms of practical therapeutic applicability. The efficiency of models in helping to identify and treat sleep-related problems early is further investigated by combining pathophysiological knowledge with empirical data.

### 4.1 Feature Importance and Clinical Interpretability

Normalized importance scores were computed across the three ensemble models to evaluate the predictive contribution of individual features. Normalization ensures comparability between algorithms with distinct internal scoring methods. Results are presented in Table 5.

**Table 5.** Normalized feature importance scores across ensemble models.

| Feature | RF | XGB | LightGBM | Mean | Std Dev |
| --- | --- | --- | --- | --- | --- |
| SystolicBP | 0.1751 | 0.1529 | 0.3008 | 0.2096 | 0.0651 |
| BMI Category | 0.1878 | 0.2511 | 0.0626 | 0.1672 | 0.0783 |
| DiastolicBP | 0.1466 | 0.1146 | 0.0603 | 0.1072 | 0.0356 |
| Occupation | 0.0939 | 0.1362 | 0.0792 | 0.1031 | 0.0242 |
| Physical Activity Level | 0.0992 | 0.1268 | 0.0540 | 0.0933 | 0.0300 |
| Person ID | 0.0801 | 0.0118 | 0.1383 | 0.0767 | 0.0517 |
| Daily Steps | 0.0594 | 0.1065 | 0.0218 | 0.0626 | 0.0347 |
| Sleep Duration | 0.0375 | 0.0263 | 0.1056 | 0.0565 | 0.0350 |
| Heart Rate | 0.0153 | 0.0367 | 0.0763 | 0.0428 | 0.0253 |
| Age | 0.0764 | 0.0075 | 0.0253 | 0.0364 | 0.0292 |
| Gender | 0.0047 | 0.0070 | 0.0643 | 0.0253 | 0.0276 |
| Quality of Sleep | 0.0132 | 0.0209 | 0.0057 | 0.0133 | 0.0062 |
| Stress Level | 0.0109 | 0.0017 | 0.0057 | 0.0061 | 0.0038 |

*Note:* RF = Random Forest; XGB = XGBoost; LightGBM = Light Gradient Boosting Machine. Feature contributions are categorized as: **Critical** (> 0.15), **High** (0.09–0.15), **Moderate** (0.05–0.09), **Low** (0.025–0.05), **Minimal** (< 0.025).

Among all predictors, systolic blood pressure (mean score = 0.2096) and BMI category (0.1672) emerged as the most critical features. LightGBM emphasized systolic blood pressure (0.3008), while XGBoost prioritized BMI (0.2511). Diastolic blood pressure, occupation, and physical activity showed high contributions, whereas sleep duration, daily steps, and heart rate had moderate effects. Demographic and subjective factors, including age, gender, sleep quality, and stress level, were consistently of minimal predictive value. Inter-model variability was greatest for BMI (std = 0.0783) and systolic blood pressure (0.0651), reflecting algorithm-specific weighting tendencies.

Consistency of feature ranking across models is reported in Table 6. Random Forest and XGBoost demonstrated moderate concordance (Kendall’s *τ* = 0.788, Spearman’s *ρ* = 0.841), with slightly weaker agreement between Random Forest and LightGBM (0.700, 0.772). XGBoost and LightGBM achieved the highest correlation (0.817, 0.863). This indicates that while all three models broadly agreed on the relative importance of key predictors, algorithm-specific weighting patterns introduced some variability.

**Table 6.** Inter-model feature ranking consistency using Kendall’s *τ* and Spearman’s *ρ*.

| Model Pair | Kendall's $\tau$ | Spearman's $\rho$ |
| --- | --- | --- |
| Random Forest vs XGBoost | 0.788 | 0.841 |
| Random Forest vs LightGBM | 0.700 | 0.772 |
| XGBoost vs LightGBM | 0.817 | 0.863 |

Clinically, the prominence of systolic blood pressure and BMI aligns with established associations between hypertension, obesity, and obstructive sleep apnea (OSA). Elevated blood pressure reflects recurrent nocturnal hypoxia and sympathetic activation in OSA, while excess adiposity promotes airway collapse during sleep. These findings highlight the capacity of non-invasive, easily measurable features to serve as actionable biomarkers for early screening in primary care.

### 4.2 Comparative Evaluation of Ensemble Classifiers

The predictive performance and generalization capability of the three ensemble models were rigorously evaluated across training, validation, and held-out test sets. Table 7 summarizes the macro-averaged accuracy and F1 scores for each data split.

**Table 7.** Accuracy and F1 scores (%) of ensemble classifiers across training, validation, and test sets (Macro-Averaged).

| Model | Accuracy |  |  | F1 Score |  |  |
| --- | --- | --- | --- | --- | --- | --- |
|  | Train | Val | Test | Train | Val | Test |
| LightGBM | 99.45 | 99.45 | **98.35** | 99.44 | 99.44 | **98.35** |
| XGBoost | 100.00 | 98.90 | 94.51 | 100.00 | 98.89 | 94.50 |
| Random Forest | 97.97 | 97.79 | 95.60 | 97.97 | 97.79 | 95.61 |

LightGBM achieved the highest test accuracy (98.35%) and F1 score (98.35%), demonstrating superior generalization. While XGBoost achieved perfect training performance (100.00% accuracy and F1), its substantially lower test scores (94.51% accuracy) and significant performance drop indicate substantial overfitting. Random Forest showed more modest but stable performance across splits.

To further assess generalization and class-wise reliability, Table 8 reports test precision, recall, and the train–test accuracy gap.

**Table 8.** Test precision, recall, and training–test accuracy gap (%) for ensemble classifiers (Macro-Averaged).

| Model | Test Precision | Test Recall | Train–Test Gap |
| --- | --- | --- | --- |
| LightGBM | **98.39** | **98.37** | **1.10** |
| Random Forest | 95.71 | 95.62 | 2.37 |
| XGBoost | 94.57 | 94.49 | 5.49 |

LightGBM demonstrated the best balance between performance and stability, achieving the highest test precision (98.39%) and recall (98.37%) with the smallest train–test accuracy gap (1.10%). In comparison, XGBoost had the largest gap (5.49%), indicating its vulnerability to overfitting despite excellent in-sample results. Random Forest demonstrated moderate generalization (gap: 2.37%) with solid but lower test metrics. These findings suggest that LightGBM is the most dependable and clinically practical model, as it combines high diagnostic accuracy with consistency on unseen data.

### 4.3 Confusion Matrix Analysis

Aggregate measures provide overall accuracy, whereas confusion matrices reveal class-specific behavior. Table 9 displays the test-set confusion matrices for all three models. Correct classifications are highlighted in green, misclassifications in red, and zero entries in light gray. Rows represent true labels, and columns represent predicted labels.

**Table 9.** Confusion matrices for ensemble classifiers on the test set.

| True ↓ / Pred → | Random Forest |  |  | XGBoost |  |  |
| --- | --- | --- | --- | --- | --- | --- |
|  | Insomnia | Sleep Apnea | Unknown | Insomnia | Sleep Apnea | Unknown |
| Insomnia | 58 | 0 | 3 | 58 | 0 | 3 |
| Sleep Apnea | 0 | 59 | 3 | 0 | 59 | 3 |
| Unknown | 2 | 0 | 57 | 4 | 0 | 55 |

| LightGBM |  |  |  |
| --- | --- | --- | --- |
| True ↓ / Pred → | Insomnia | Sleep Apnea | Unknown |
| Insomnia | 59 | 0 | 2 |
| Sleep Apnea | 0 | 61 | 1 |
| Unknown | 0 | 0 | 59 |

#### Interpretation

Random Forest correctly classified most cases but misidentified three insomnia patients and three sleep apnea patients as “Unknown,” along with two “Unknown” cases labeled as insomnia. XGBoost showed a similar pattern but with slightly higher error, misclassifying four “Unknown” cases as insomnia. In contrast, LightGBM produced the most accurate predictions, with only two insomnia cases and one sleep apnea case mislabeled as “Unknown,” achieving perfect separation for the “Unknown” class otherwise.

Across all models, no instances of sleep apnea were falsely categorized as insomnia, ensuring high clinical safety by avoiding false negatives for a life-threatening disorder. The consistent misclassification of insomnia as “Unknown” indicates overlapping symptom profiles that the current feature set cannot fully resolve. Incorporating physiological measures such as oxygen saturation or respiratory effort could further enhance discriminatory power.

Overall, LightGBM demonstrated the strongest diagnostic reliability, minimizing false positives while maintaining excellent recall for high-risk conditions, making it the most clinically dependable candidate for deployment.

### 4.4 Comparison with Existing Literature

To better understand the findings, the Random Forest, XGBoost, and LightGBM models are compared to previous work on sleep disorder categorization (Table 10). Rahman et al. (2025) reported 97.33% accuracy using gradient boosting ensembles, while Alshammari (2024) obtained 92.92% with artificial neural networks (ANN) and standard machine learning techniques.

**Table 10.** Comparative performance of sleep disorder classification models.

| Study / Model | Accuracy (%) | Precision (%) | Recall (%) | F1-score (%) |
| --- | --- | --- | --- | --- |
| This Study | 98.35 | 98.39 | 98.37 | 98.35 |
| Rahman et al. (2025) (Gradient Boosting) | 97.33 | 97.33 | 97.33 | 97.33 |
| Alshammari (2024) (ANN) | 92.92 | 92.01 | 93.80 | 91.93 |

While both this study and that of Rahman et al. (2025) employed class balancing through resampling approaches, such as SMOTE, the current work advances the field in two crucial areas. First, hyperparameters were tweaked individually for each model rather than universally, ensuring that optimization matched the algorithms’ inductive features. Second, clinical dependability was highlighted, and the LightGBM model achieved a near-perfect recall for sleep apnea, thus reducing the likelihood of false negatives in a high-risk medical setting.

In this research, the LightGBM model outperformed the model reported by Rahman et al. (2025), reaching 98.39% precision and 98.37% recall, with a train-test gap of just 1.10%, suggesting outstanding generalization and stability. Random Forest and XGBoost also performed well, confirming the efficacy of tree-based ensembles for structured medical datasets. In comparison, the ANN-based technique of Alshammari (2024) yielded significantly lower accuracy, highlighting the ongoing benefits of ensemble approaches when interpretability, robustness, and clinical safety are prioritized.

These findings demonstrate that even with similar preprocessing procedures, careful model selection and rigorous tuning are necessary to achieve cutting-edge clinically significant results.

### 4.5 Deployment of the Optimal Model

Following a thorough study, LightGBM was chosen as the best model for deployment due to its excellent test performance, small training-test gap, and strong clinical safety profile. The final trained model was serialized using Python’s `joblib` module, allowing efficient inference without retraining.

A lightweight REST API was developed using the `Flask` framework to handle prediction queries. The front-end, designed to provide physicians with an intuitive and responsive user experience, was implemented with standard web technologies, including HTML, CSS, and JavaScript. Incoming patient data are preprocessed prior to prediction to align with the model’s training schema, and results are returned immediately to support rapid clinical decision making.

Figure 2 illustrates the deployed interface, showing how clinicians can input key patient parameters and receive immediate, interpretable predictions.

**Figure 2.**
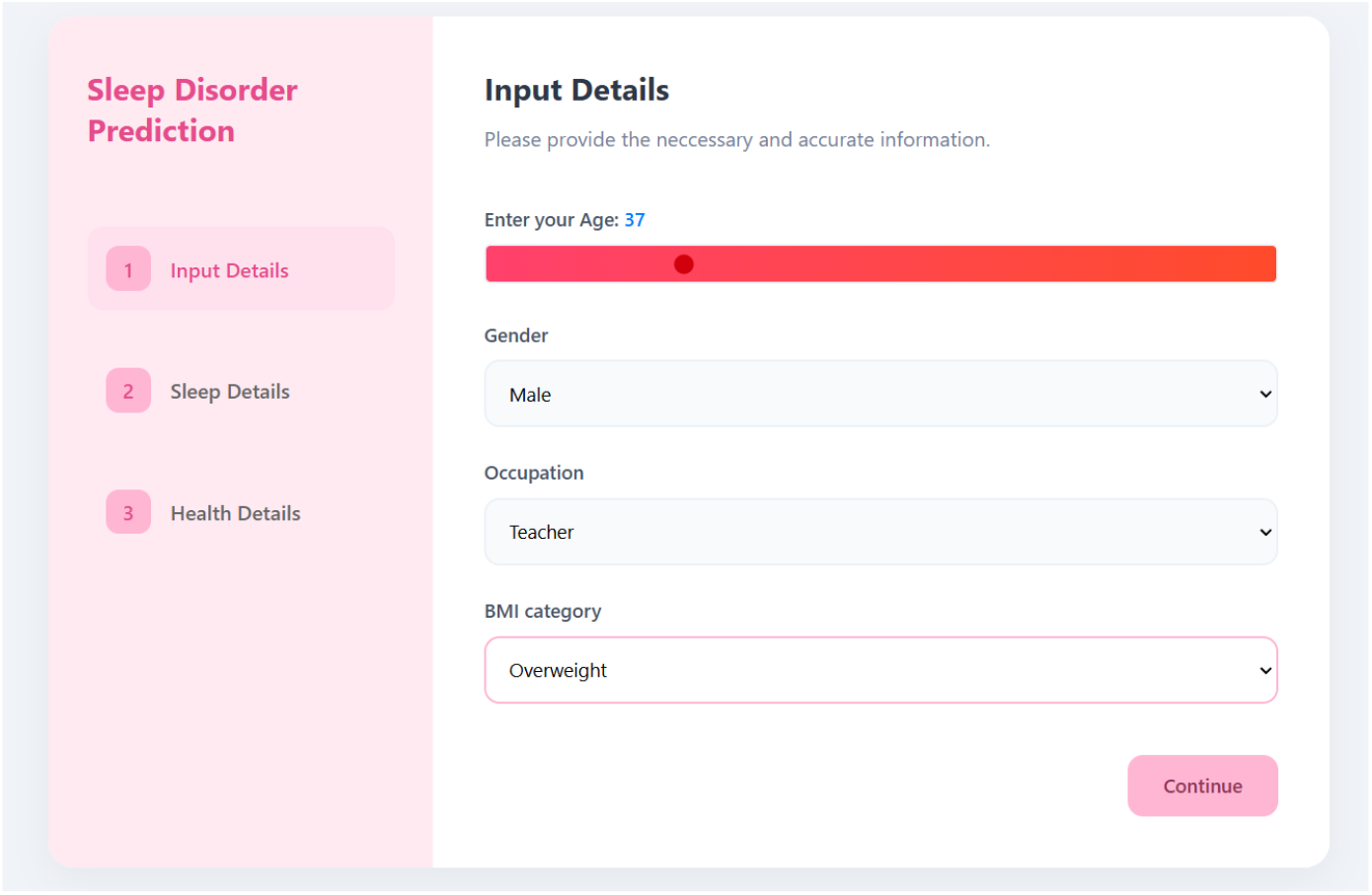
User interface of the deployed Random Forest model.

## 5 CONCLUSION

This paper presents an efficient machine learning framework for multi-class sleep disorder classification using only the original attributes of the Sleep Health and Lifestyle Dataset, without reliance on engineered features. LightGBM emerged as the best-performing model, achieving a test precision of 98.39%, recall of 98.37%, and a minimal train–test gap of 1.10% through SMOTEENN-based resampling and systematic hyperparameter tuning. In particular, the model maintained high recall for sleep apnea, thus minimizing false negatives, a critical requirement for safe clinical screening.

Compared with other ensemble approaches in this study, LightGBM consistently outperformed both Random Forest and XGBoost, delivering higher test precision, stronger generalization, and better resilience to overfitting. Compared with previous work, including Rahman et al. (2025) who reported slightly lower accuracy with gradient boosting, the LightGBM model in this work demonstrates superior performance, improved generalization, and greater robustness against overfitting.

The framework has been operationalized as a Flask-based REST API, enabling real-time predictions from structured patient inputs and offering scalable integration into clinics, telemedicine platforms, and low-resource environments. Future directions include large-scale external validation, incorporation of physiological signals from wearable devices, and the adoption of explainable AI techniques to enhance clinician confidence and model transparency.

Overall, the findings confirm that carefully optimized gradient boost ensembles, particularly LightGBM, can deliver accurate, safe, and deployable solutions for the classification of sleep disorders, supporting early diagnosis and improved access to sleep health care.

## Data Availability

full available

